# Evolution of case fatality rates in the second wave of coronavirus in England: effects of false positives, a Variant of Concern and vaccination

**DOI:** 10.1101/2021.04.14.21255385

**Authors:** James A Ackland, Graeme J Ackland, David J Wallace

**Affiliations:** Department of Psychology, University of Cambridge, CB2 3EB; School of Physics and Astronomy, University of Edinburgh, EH9 3FD; International Centre for Mathematical Sciences, Edinburgh, EH8 9BT

## Abstract

1

**Objective:** To track the statistical case fatality rate (CFR) in the second wave of the UK coronavirus outbreak, and to understand its variations over time.

**Design:** Publicly available UK government data and clinical evidence on the time between first positive PCR test and death are used to determine the relationships between reported cases and deaths, according to age groups and across regions in England.

**Main Outcome Measures:** Estimates of case fatality rates and their variations over time.

**Results:** Throughout October and November 2020, deaths in England can be broadly understood in terms of CFRs which are approximately constant over time. The same CFRs prove a poor predictor of deaths when applied back to September, when prevalence of the virus was comparatively low, suggesting that the potential effect of false positive tests needs to be taken into account. Similarly, increasing CFRs are needed to match cases to deaths when projecting the model forwards into December. The growth of the S gene dropout VOC in December occurs too late to explain this increase in CFR alone, but at 33% increased mortality, it can explain the peak in deaths in January. Seasonal effects could be in part responsible for the early December increase in CFR, and if so, the estimate of increased mortality would be reduced. There is also evidence that the prevalence of B.1.1.7 may have been slower amongst older age groups, and if this is a factor, then 33% could be an underestimate of mortality. From the second half of January, the CFRs for older age groups show a marked decline. Since the fraction of the VOC has not decreased, this decline is likely to be the result of the rollout of vaccination. However, due to the rapidly decreasing nature of the raw cases, any imprecisions in the time-to-death distribution are magnified in this time period, rendering estimates of vaccination’s effect less precise.

**Conclusions:** The relationship between cases and deaths, even when controlling for age, is not static through the second wave of coronavirus in England. An apparently anomalous low case-fatality ratio in September can be accounted for by a 0.4% false-positive fraction. The rapid growth in CFR in December can be understood in part in terms of a more deadly new variant B.1.1.7, while a decline in January correlates with vaccine roll-out, suggesting that vaccine reduce the severity of infection, as well as the risk.

**Summary Box:** *What is already known on this topic:* The case fatality rate (CFR) is a useful measure which enables one to estimate future deaths based on current infections. In England, there was a surge in Covid-19 CFR around the beginning of December.

*What the study adds:* Using it, we monitor the case-fatality rate across time, region and age group from publicly available case data. This quantity is related to the lethality of the virus. It shows a sharp increase in December 2020, which parallels the spread of the B.1.1.7 variant. The January peak in actual deaths matches that predicted by cases if B.1.1.7 is about 33% more deadly; this estimate would be lower if there is a seasonal effect on deaths, and higher if at the peak the variant was less pervasive amongst older age groups. A steady drop in CFR from January suggests that vaccination not only reduces transmission but also the risk of serious illness among those infected. It is notable that these conclusions are reached with publicly available data independent of clinical studies.

## 2 Introduction

The ongoing coronavirus pandemic has emphasized the importance of practical epidemiological modelling and surveillance. Quantities which are straightforward to define in theory, can be extremely tricky to evaluate in the face of incomplete, confidential and noisy data. In a fast-developing situation, it becomes essential to build theories around what data is available, rather than the data one would like to have.

Patient and case data exemplify the point. In the UK, such data are not publicly available, and in many countries may not be available at all. Vaccines such as Pfizer require careful handling and cold chains, so there is serious risk of substandard doses, and falsified medicine is a massive problem in many parts of the world. So a reliable method of evaluating real-world vaccine efficacy based only on epidemiological data, and robust against incompleteness of that data, is very important.

The Infection-fatality ratio (IFR), which determines how deadly the disease is, is a central parameter in most epidemic modelling, but it has proved difficult to determine empirically. The very high fraction of asymptomatic cases means that the true number of coronavirus infections may never be known. In practice we need to work with the available data which comes from testing, and the ratio of positive tests (“cases”) to coronavirus-linked deaths. This is the case-fatality ratio (CFR).

The CFR has a number of useful properties not least because it is precisely defined by publicly available data [1]. Moreover, because both cases and deaths rise and fall together with the prevalence of the virus, the CFR is insensitive to case numbers. Changes in the CFR will therefore be due to other factors such as virulence of variants, or effectiveness of vaccines in reducing deaths. Thus CFR is a complementary measure to, for example, the R-number which measures infection rates.

The heat maps shown in Figure 1 illustrate how a useful CFR is not a simple ratio. In addition to the scaling factor between cases and deaths, it is immediately obvious that the peak in deaths is shifted forward in time and to older age groups. In a previous paper [2] we built a simple predictive model for deaths based on the CFR, where the case data is *weighted* for age group, *scaled* for survival probability and *shifted* for the time between first positive test and death. The weighted-shifted-scaled model (hereinafter WSS) is based on the idea of a constant CFR.

**Figure 1:**
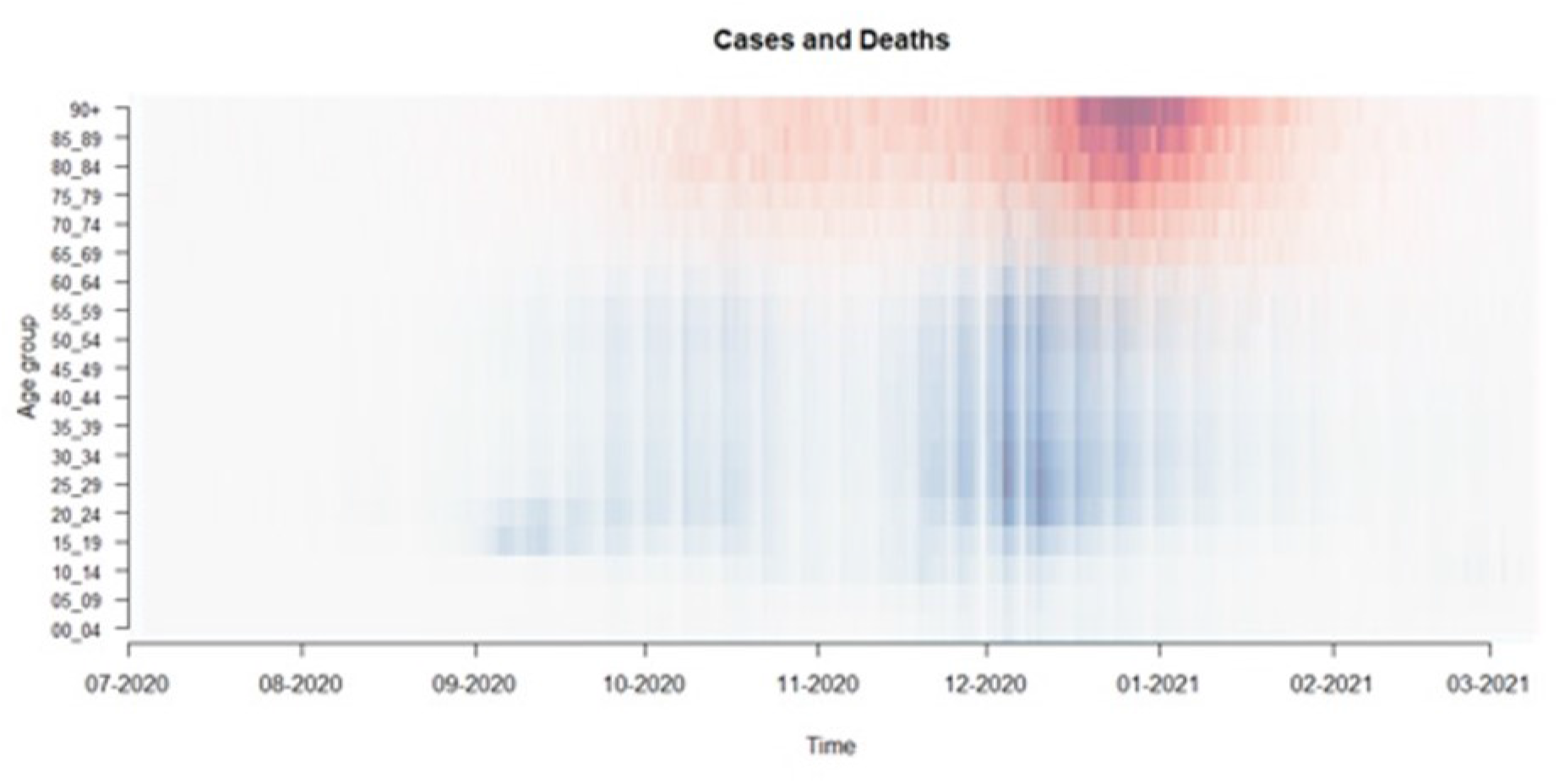
Overlaid heat maps for cases (blue) and deaths (red) by age group 0-4 to 90+ and by date from October 2020. The deaths may be related to cases by Weighting by age, Shifting forward in time and Scaling by a case fatality ratio. The ‘weekend effect’ in cases reporting is visible in vertical stripes, while no such effect is noticeable in deaths reporting.

Although the age weighting of CFR is extreme in COVID, it has surprisingly little signal in the overall data. This is because it averages out when the infection is evenly spread throughout the age groups. Age-differences in CFR only affect overall CFR when there are also age differences in case rates. Heat maps like Figure 1 show that during the first wave in the UK in early 2020, infection arrived in middle-age groups, but after lockdown it became stronger in the older age groups, particularly in care homes. This skew was so significant that, despite the CFR in all age groups being higher for men than women, overall more women were dying. In September, the resurgence of infection began in young adults, and gradually shifted upwards through the age ranges, with a pronounced dip in the more recently retired 65-75 age range.

In [2], the age-weighting was taken from a CDC study [3] (updated now as of Feb. 18 2021), and assumed to be constant. Parameters were then estimated to describe: 1) A “shift” representing the average number of days from specimen date of test to death, and 2) A ‘’scale’’ factor (the CFR) converting the relative likelihood of death to the absolute likelihood of death. Analysis of the relationship between coronavirus cases and subsequent deaths in England [2] revealed an excellent predictive power for the WSS from October through November based on constant shift and scale parameters. Specifically, these ‘shift and scale’ factors were fixed by matching cases-predicted deaths to actual deaths at the 21^st^ November peak, which was just over 400 deaths per day. The analysis used 7-day averaging, to avoid the ‘weekend effect’ prevalent in the reporting of cases by specimen date.

We then found that in December 2020 this well-fitted WSS model abruptly started to underpredict the ratio of actual deaths to deaths predicted from the number of cases (CFR) failing firstly in southern England, and later in mid-December in Yorkshire and Scotland. This was the first published evidence suggesting that the new variant B.1.1.7 [4] might be not only more infectious, but also more lethal, a conclusion also reported by SAGE and confirmed by a number of subsequent publications using confidential case data [5,6]. Another recent clinical study [7] rules out increased lethality of B.1.1.7 vs non-B.1.1.7 above 85%.

The CFR anomalies identified by WSS [2] do not prove causation. The PyRoss collaboration [8] show that Bayesian inference based on various compartment models can be used to quantify the likelihood of causation. They argue that while the new variant(s) may be one contributing cause of a large increase in fatality rate in autumn 2020, other factors, such as seasonality, or pressure on health services, are likely also to have contributed.

In this paper, we present a continuation of our analysis covering the period of vaccine rollout. The drug trials for vaccines looked at prevention of detectable infection, finding typically an 80-90% efficacy. The number of COVID deaths in the trials was too small to draw conclusions about CFR. As explained above, the WSS model should be insensitive to any changes in infection rates, so any vaccination-related reduction in the WSS predictions would indicate that vaccination makes infections *less deadly*, in addition to the known effect of less likely.

The WSS model may be regarded as an algorithm for prediction of deaths based on case data, via a fixed CFR. Alternately, it can be used with contemporary deaths data to generate time-varying CFR, which can give insight into the evolution of the virus. It is this second approach which is utilized here.

## 3 Improving the WSS Method

In this paper, we replace the naive ‘shift’ assumption used in [2] by a distribution of time to death. Analysing cases and deaths data for England for each of the age groups enables us to determine CFRs for each age group, without depending on the values inferred from the CDC study [3] in [2], which may no longer apply to the new variants and circumstances of the pandemic in England.

### 3.1 Distributions of time to death

To identify an appropriate distribution to improve the ‘shift’ assumption, we have reviewed six studies reporting time to death distributions [9-14]. The distributions found in these studies share the characteristic of being long-tailed, and are variously fitted by lognormal, gamma or Weibull distributions (Figure 2).

**Figure 2:**
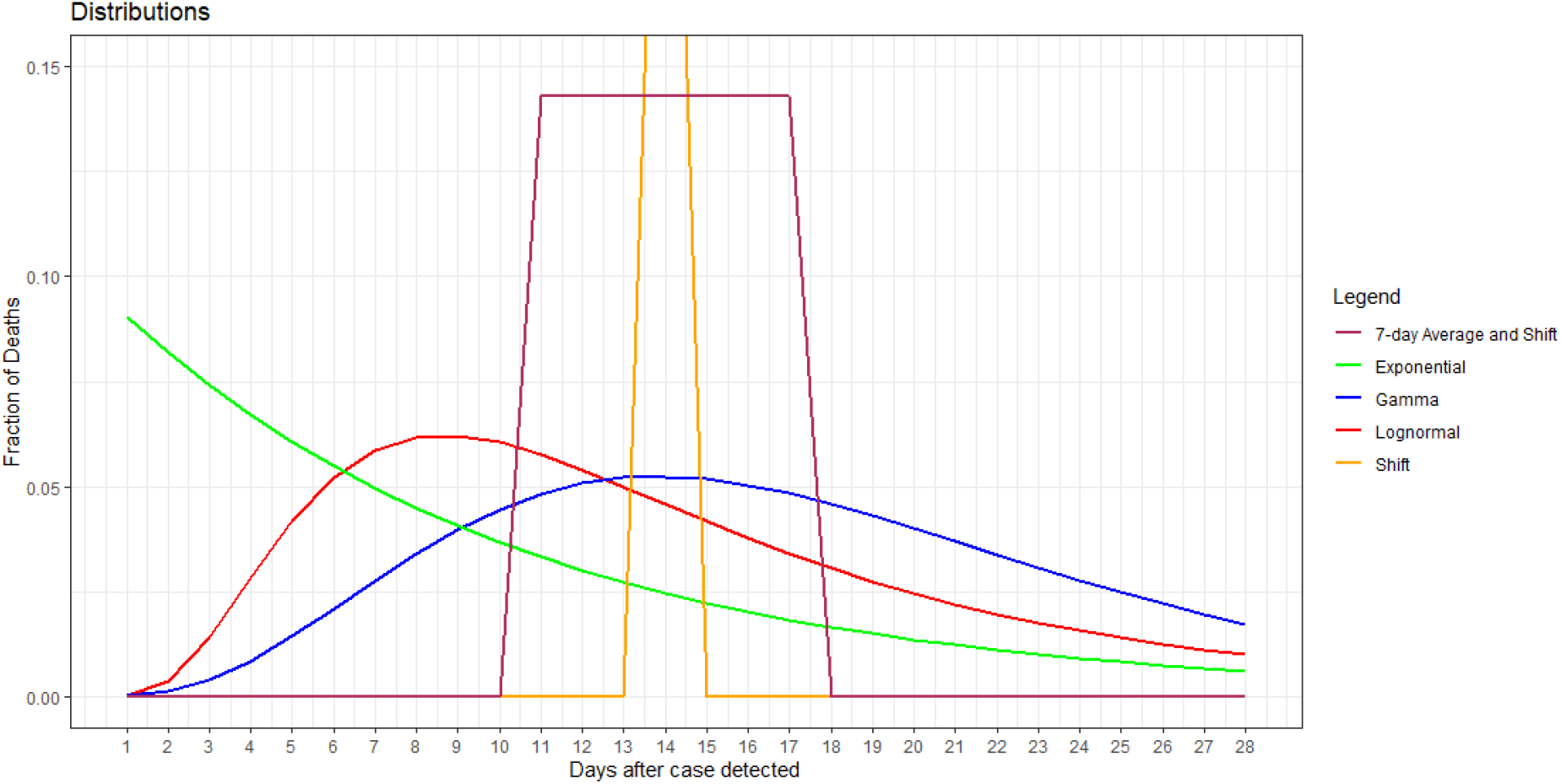
Various different distributions of time to death. The prediction for deaths is a convolution of this function with the case data. For illustration the mean of each distribution is set to 15 days following Verity et al. [9], and the standard deviation of the gamma and lognormal are set to 10.26. The exponential distribution is implicit in differential equation treatments such as Kermack and McKendrick’s SIR model. All distributions are truncated at 28 days to meet the UK definition of “COVID death”.

Some of these studies report distributions for time to death following hospital admission, which would not be appropriate to describe the distribution of first positive tests to death. A second criterion for whether a distribution is appropriate for our work is that the cumulative value within 28 days compared with that for 60 days should be consistent with England data for the ratio of deaths within 28 days to deaths within 60 days (0.91). There are two clinical studies for the distribution for onset of symptoms to death which meet this criterion: Verity et al [9] and Hawryluk et al [13].

In order to test the validity of our approach, we analyse data using a lognormal distribution with the mean and standard deviation from [13] and a gamma distribution based on the parameters from [9]. The lognormal distribution parameters suggested in [13] are µ = 2.53 and s = 0.613, corresponding to an average time to death and standard deviation of 15.2 and 10.26 days respectively. The gamma distributions parameters suggested in [9] are α = 4.45 and β = 4,00, corresponding to an average time to death and standard deviation of 17.8 and 8.44 days respectively. Since English COVID data is counted by deaths within 28 days, we use the discrete values for days = 1 to 28 from these distributions, thus allowing average times to death to depend on age, and for deaths to occur uncounted beyond 28 day.

The long tails of the gamma and exponential distributions largely eliminate the ‘weekend effect’ in cases, and so obviate the need for seven-day averages used in [2].

Calculations were performed independently in R and in Excel, providing an element of code verification.

### 3.2 Mortality rates by age

Mortality rates in COVID vary strongly by age, so it is essential to weight cases from each age group separately (difference between dashed and solid curves in figure 6). Previously this was achieved using data inferred from a CDC study based on mid-2020 US data [3]. Instead, age weights are determined by the UK data [1]. We condition the model on the period of October and November. This period is suitable for three reasons. First, both cases and deaths were relatively high, ensuring a sufficiently high quality of data to estimate age weights for the ten oldest age groups. Second, the period includes both increasing and decreasing phases of cases. Third, neither vaccination nor variant of concern B.1.1.7. were significant factors at this time, both of which may have affected the relationship between cases and deaths within age groups. Age weights are calculated as the regression line between deaths and gamma-distributed cases, and can be interpreted as the percentage of cases expected to die (see Table 1 and Fig. 4).

**Table 1.** Estimated age weights conditioned on October and November data, with 95% confidence intervals. For future reference, we note that these weights do not take the effect of false positives into account.

| Age Group | 45_49 | 50_54 | 55_59 | 60_64 | 65_69 | 70_74 | 75_79 | 80_84 | 85_89 | 90+ |
| --- | --- | --- | --- | --- | --- | --- | --- | --- | --- | --- |
| Weight | 0.22% | 0.36% | 0.75% | 1.18% | 4.51% | 8.18% | 14.12% | 22.21% | 29.97% | 33.63% |
| 95%CI Lower | 0.12% | 0.26% | 0.57% | 0.93% | 3.93% | 7.33% | 12.88% | 20.23% | 27.70% | 31.23% |
| 95%CI Upper | 0.32% | 0.45% | 0.93% | 1.43% | 5.09% | 9.03% | 15.35% | 24.20% | 32.25% | 36.03% |

These weights incorporate CFR at the levels seen in October and November, and replace both the CDC ‘weight’ and ‘scale’ parameters used in our original model [2].

### 3.3 Testing and parameterizing the WSS model

The model predictions compared with actual deaths are shown the table below and in Figure 3. The linear model correlation of cases-expected deaths and actual deaths is good for the national data both for October and November and for the full time-series. The good fit in Autumn months is unsurprising given these were the dates on which age weights were conditioned.

**Figure 3.**
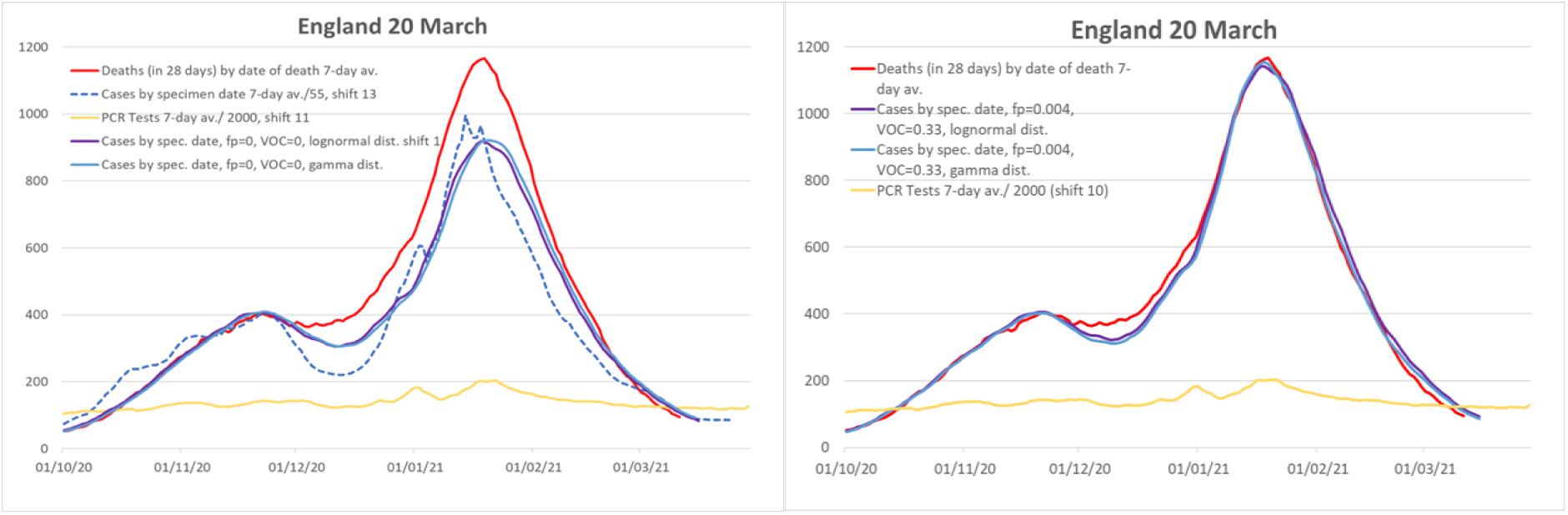
Actual and cases-expected deaths up to March 20th: Graphs show reported deaths (red), expected deaths from simple scale-and shift of cases, and our model including age-weights and lognormal/gaussian distribution. The lefthand graph, shows the WSS predictions assuming no false positives and no more lethal B.1.1.7, the righthand graph, which includes effects of VOC and false positives from section 4 and is included here for ease of comparison. The yellow lines show the number of PCR tests reported, which is roughly constant: at this level of testing, there is nothing to suggest that the number of tests affects the number of cases (the so-called casedemic).

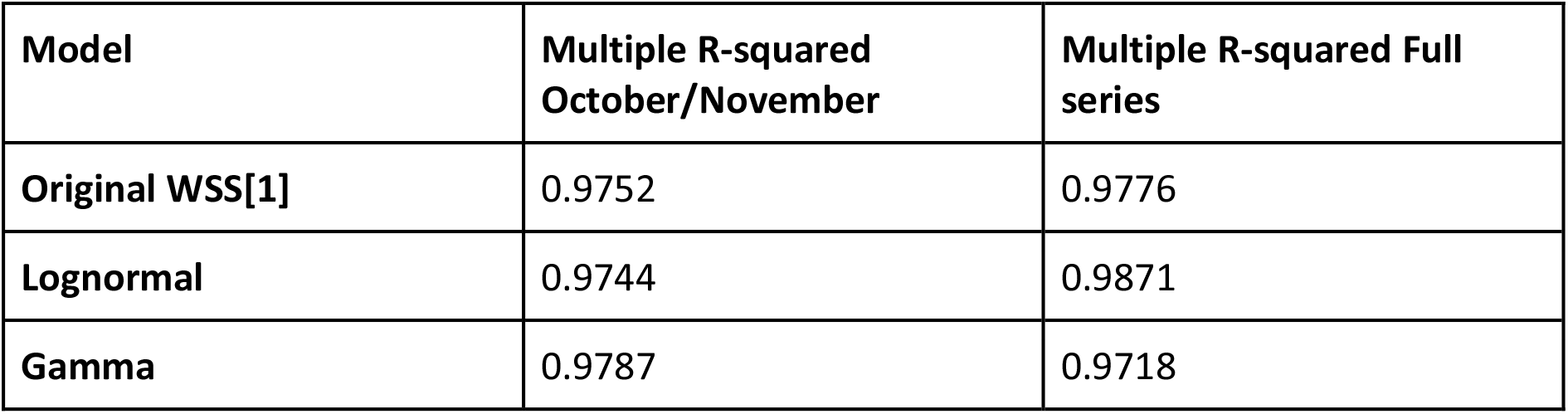

| Model | Multiple R-squared<br>October/November | Multiple R-squared Full<br>series |
| --- | --- | --- |
| Original WSS[1] | 0.9752 | 0.9776 |
| Lognormal | 0.9744 | 0.9871 |
| Gamma | 0.9787 | 0.9718 |

Although the linear model correction is high, Figure 3 shows the surge in deaths during December, noted in [2].

We initially assumed that all weight, scale and shift parameters are constant in time: we can test this by calculating the day-by-day value of case-fatality rates over time *t*, defined by:

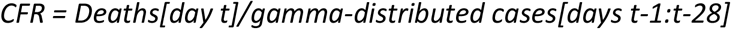

Such CFRs can be defined separately for any stratification for which breakdowns of both cases and deaths data are available, e.g. by age group or by region.

To further validate the trends in Figures 3 and 4. we restratify the data across various regions rather than age (Figure 5). If the variations we see are indeed effects rather than statistical artefacts, then they ought to persist in any arbitrary divisions of the data - region is chosen because the base-rate CFR discrepancies are also of interest. The transferability of the WSS model from National to Regional level gives us confidence that the model is a reliable one.

**Figure 4.**
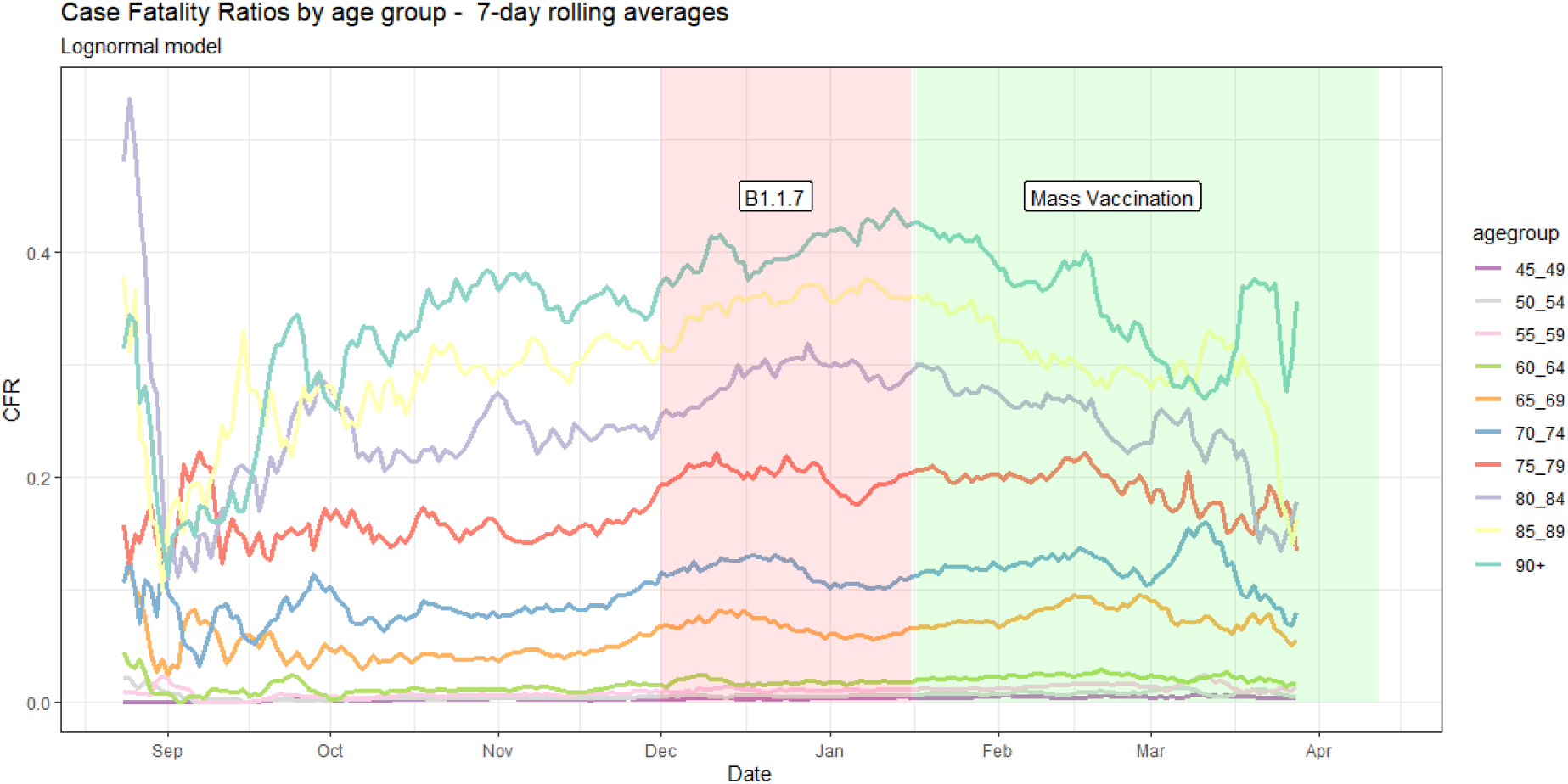
CFRs by date and age group,. The main phase of variant B.1.1.7 growth is highlighted in red, while the period during which vaccination can be expected to have a meaningful effect (14 days after 2% of the population had received their first dose), is highlighted in green.

**Figure 5.**
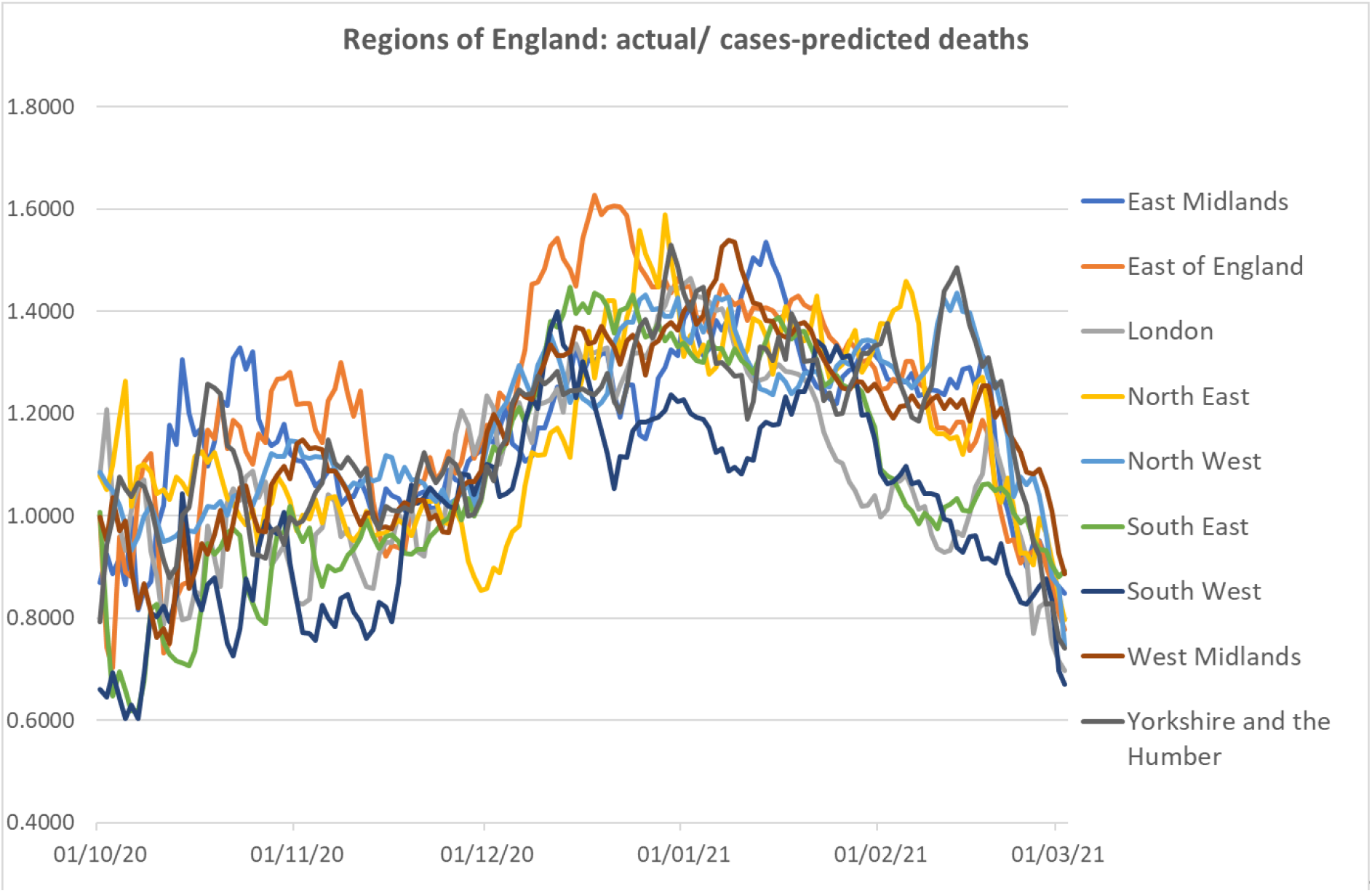
shows the data for the various English regions, comparing predictions of WSS parameterised using CFR national averages with the actual death data. This can be thought of as a regional “weighting” for CFR, analogous to age weighting. Although the data is noisy, the shape of the time-series mirrors that of Fig 7. This provides additional evidence that these trends are not noise. We note in passing that there are substantial regional differences in CFR which persist over time, as shown in Table 2

As already noted in [1], CFRs rise abruptly in December, and fall from January onwards. These trends are systematic across all age groups and all regions, and so cannot be attributed to noise.

**Table 2.** Regional variations in case-fatality ratios and standard error (σ /√N). Averages are across all age groups, standard error based on daily fluctuations. Also shown is the increase in CFR between October-November and the December-January plateaux.

| Region | England | East Midlands | East | London | North East | North West | South East | South West | West Midlands | Yorkshire & Humberside |
| --- | --- | --- | --- | --- | --- | --- | --- | --- | --- | --- |
| CFR % difference | 0 | +7.8<br>(1.3) | +7.5<br>(1.7) | -2.8<br>(1.5) | +2.4<br>(0.8) | +4.9<br>(0.9) | -11.5<br>(1.6) | -14.2<br>(1.6) | -0.6<br>(1.3) | +4.2<br>(1.1) |
| B.1.1.7 CFR increase | 33% | 20% | 32% | 31% | 26% | 24% | 48% | 40% | 34% | 24% |

Table 2 shows how the regional CFRs highlight significant differences across England, with generally better outcomes in Southern regions than Northern ones. The data cannot reveal the cause for this. They also show that the arrival on B.1.1.7 had a significant effect across all regions. Interestingly, Figure 5 shows that the regional differences became quite small during December, when the southern areas were more heavily affected by B.1.1.7 which appears to override their long-term advantage. Once B.1.1.7 is established everywhere, the regional differences reappear.

## 4. Results

### 4.1 Detecting false positives

We observe that the reported number of deaths in September [2] is significantly lower than the simple WSS prediction. A discrepancy should be expected due to small-number statistics, but we found that the lower CFR is visible independently in each age group albeit with even noisier statistics. Were the model’s overprediction purely noise, it should not survive stratification. We hypothesise that this apparently-reduced CFR is in fact a function of a comparatively high false-positive rate in testing during this period, with no concomitant inflation in deaths.

The relatively small number of cases in September (2,000 rising to 10,000 per day) along with the rather large number of tests (c. 150,000 to more than 200,000 or more per day) enables one to estimate the false positive fraction for PCR tests [16,17]. There are two types of cause for false positives: errors in the test and reporting itself, which we expect to be a small constant fraction, and errors from sample contamination which will increase with the amount of true positives.

When case numbers are lower, false positives will form a higher fraction of total case numbers. To account for this we can deduce the false positives using the relation…

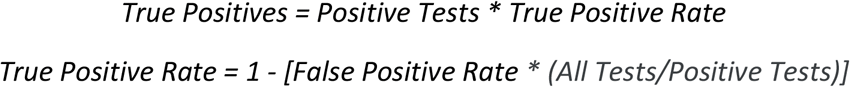

Estimating the effect of false positive tests is complicated by practical factors.

First, the number of false positives depends on the number of tests conducted. Until 20 September 2020, data on the number of tests are publicly available only for the UK as a whole. We estimate the number for England by using the average proportion of UK tests that occurred in England during the period 21 September to 5 November (0.867).

Second, although data on tests are available by date of test, the number of tests is available only by published date. These measures are out of phase and are an unfortunate illustration of the issues arising from a weekend effect on test activities and reporting (see Fig. 6). We ameliorate this effect by using the test data announced two days after the specimen date. We also tested using a one-day shift, and 7-day averaging of tests data; the results are not changed materially.

**Figure 6:**
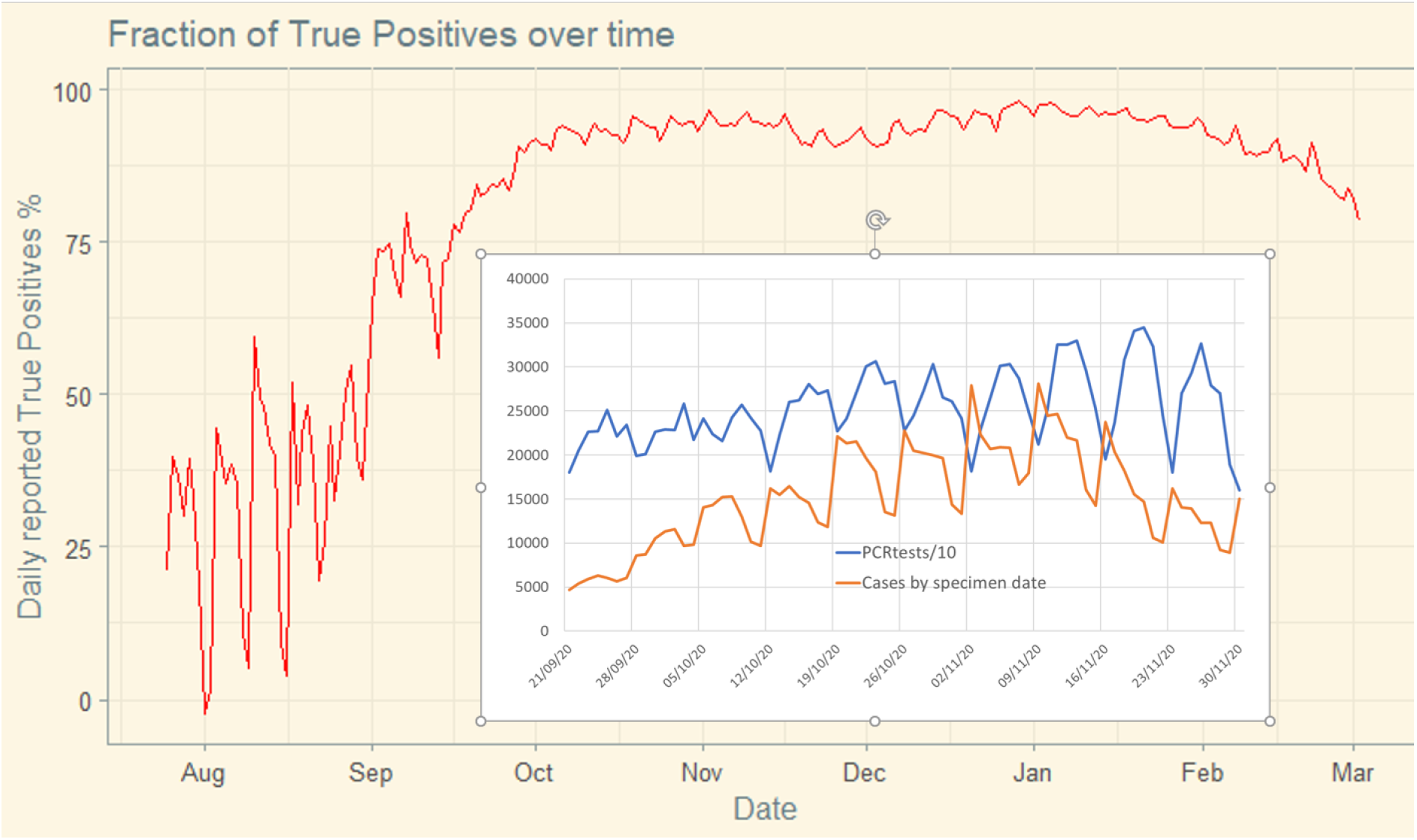
Our estimate of the proportion of positive PCR tests that can be considered true positives each day. Note the extreme volatility and low average validity in the early phase of the data. This chart uses the false positive fraction 0.004: inset show time offset between test reports and case data leading to spurious weekly oscillation.

Third, since we analyse cases and deaths by age group, we need the number of tests carried out for each age group. These data are not available publicly, so instead we use the reported PCR tests for each day multiplied by the fraction of positive tests for that age group. This is a significant assumption, but is being used in what for most of the data is a small correction.

Finally, there has been a recent increase in lateral flow tests, which are known to have very high false positive rates. For our purposes these are ignored except when backed up by a subsequent PCR test. This introduces a reporting delay.

The number of new daily positive tests was around 2000 at the beginning of September and increased to 10,000 by the end of month. PCR tests conducted daily rose from around 160,000 to 220,000 in the same period. The linear fits to these data suggest a false positive fraction between 0.004 to 0.005. Other estimates for this number vary. [15] suggests a rather larger number. The small number of positive cases in ONS data from summer 2020, implies the total positive figure, an upper bound on the false positives, was just 0.00005[17,18]. The cause of false-positives in PCR tests is not known, but if sample contamination were important then it would explain how false-positive rates could rise as total rates increase.

Figure 9 shows the fraction of true positives if a fraction 0.004 of PCR tests are false positives. This rate has little effect when cases are high, but in September most positives tests are likely to be false, and, increasingly, in March: with some 250,000 PCR conducted daily, and the average number of new cases falling to around 5000, false positives of around 0.4% represent around 20% of new cases reported.

If the low summer value of 0.00005 continued into September, we would need to seek another explanation for why CFRs in September were so much lower than CFRs in October and November. Deterioration of treatment quality seems unlikely, an alternative possibility may be that the B.1.177 “Spanish” variant, which is slightly more infectious, and became dominant during September [19], is significantly more deadly. Evidence for this is weak, and we have been unable to find any clinical data to support this. At the time of writing, case numbers are again low and the false positive rate is again significant.

### 4.2 Detecting effects from the VOC B.1.1.7

Data from ONS and COG-UK [19,20,21] showing that the slightly more infection B.1.177 (“Spanish”) variant became dominant during September and October, and the significant more infectious B.1.1.7 (“Kent”) took over in December. Separate R-numbers for these variants show that the original variant has been under control since November, that B.1.1.7 was responsible for the Christmas “peak” and was only brought under control at the time of the January national lockdown.

We hypothesise that the abrupt increase in CFR in early December [1] is due to the variant of concern B.1.1.7. To test and evaluate this, we will adjust the “scale” factor in the WSS with a term depending on the fraction of B.1.1.7 cases. The B.1.1.7 prevalence varies with time, age and region so there are several independent data streams against which to test the hypothesis.

#### 4.2.1 Spread of the Variant B.1.1.7

If one considers two competing variants, conferring mutual immunity, and with different infectivities favouring the new variant by a factor b, the rates of growth will be

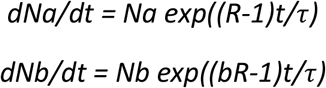

Where Na, Nb are case numbers for the two variants, R is the “R-number” of variant a. *t* is the time, *τ*is the generation time. *t=0* can be chosen arbitrarily, the choice determines *Na(0)* and *Nb(0)*. Ignoring effects from immunity, a little algebra shows that the fraction of b is expected to grow according to a logistic relation. Defining the ratio of variants as *A*(*t*) = *Nb*(*t*)/*Na*(*t*) we find

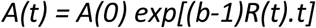

This is not quite a logistic relationship, because of the *R(t)* term, but in practice variations in *R(t)* are small enough that the data is close to logistic.

Data tor tracking the Variant of Concern (VOC), for England come from the infection survey carried out by the Office for National Statistics (ONS) [18]. This survey monitors the prevalence of coronavirus in a representative fixed cohort of people. In general, it gives an alternative measure of the number of cases, free from the uncontrolled and unrepresentative testing in the UK Government data. Similar data from Public Health Scotland (PHS) was obtained by Freedom of Information [22]. Finally, data from COG-UK consortium genomics survey is provided via the Sanger Institute

Before 31 October, ONS reported for each day the modelled percentage testing positive. From 31 October, this number was broken down into two categories: ‘new variant compatible’ and ‘other’. Unfortunately, when cases started to drop, the viral content in many samples became too small for the variant to be detectable, and the reported B.1.1.7 fraction, implausible, began to drop. This was because B.1.1.7 cases with low viral load were reported as ‘other’. Only since 13 December, ONS has reported under three categories: new variant compatible, other, and virus too low for variant to be identifiable.

The PHS data for Scotland show the fraction of VOC over the total. ONS data for Scotland show the fraction of ‘new variant compatible’ for the two cases where the denominator is the total %, and where it is the sum of ‘new variant compatible’ and ‘other’, i.e. disregarding ‘too low to be identifiable’. There is striking agreement between the PHS data and ONS data when the % ‘too low to be identifiable’ is assumed to have the same fraction of VOC as the identifiable samples. Data collected by the Sanger Institute’s Genomic survey team shows a very similar behaviour, also identifying the rise of the B.1.177 variant [19].

The growth of new variant in Scotland appears to follow the same logistic pattern as in England, with a delay of 3-4 weeks. Between 25 November and 2 January, the ONS England B.1.1.7 fraction is similar to the PHS Scotland data shifted by 27 days, and with ONS Scotland data by 28 days. For the former, the standard deviation by data point is 3%, which is largely accounted for by the daily variance of the PHS data.

On this basis, our estimate for effective % VOC for England and English regions is to use the available post-13 December to fit a logistic relation for each region.

The England data after 13 December and the Scotland data are well fitted by the logistic function *1/[1 + exp k(t-t*_*0*_)] where the units of *t* are days, *t*_*0*_ occurs on 9 December, and *k* = 0.06. Noting that *k=(b-1)R/τ*, and taking *b=1*.*33; R=1* this gives an independent estimate of the generation time of *τ=5* days.

We note that a working paper [23] to Sage ‘includes a correction for the probability over time that a specimen with s-gene target failure is the VOC’. The nature of the correction is not specified.

A further issue is that the headline figures in the ONS survey are not for *new* cases; if subsequent tests within the cohort also return positives, they are included. Thus the actual increase in B.1.1.7 occurred earlier and faster than the ONS data suggest.

#### 4.2.2 Including the effect of the VOC

Including the effects of VOC with increased mortality, and prevalence given by our logistic fit, is a straightforward one-parameter embellishment to WSS. As we noted in [1], the fit to data suggests a 33% increase in mortality, but the data suggests a faster onset and a detailed Bayesian inference study [8] suggested a much higher value. Fig 8 shows that the value of 33% gives agreement at and around the peak of deaths in January and with more recent data. This 33% value is consistent with first estimates from clinical studies [7, 23], but lower than a recent epidemiological study [24]. It also suggests that the increased CFR is similar to the increased infectivity: these properties are cumulative, which makes the B.1.1.7 variant much more dangerous.

**Figure 7.**
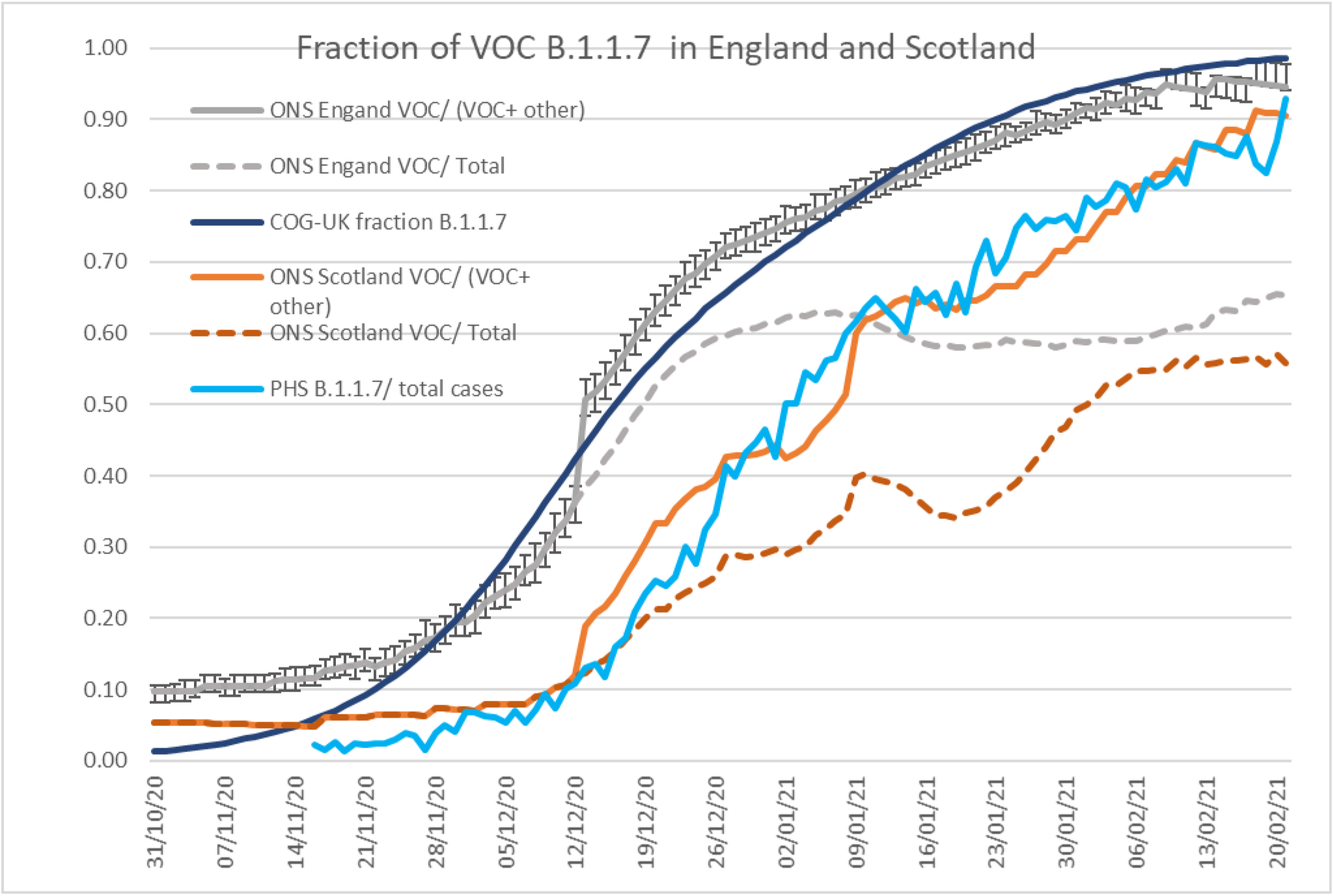
Fraction of new variant from ONS [18], COG-UK [19] and Public Health Scotland [22] data. Agreement for both the England and Scotland data is good only if the % ‘too low to be identifiable’ is excluded from the denominator. The dashed lines show the problem of including that fraction, i.e. assuming that samples which are not identifiable as B.1.1.7 are not B.1.1.7: the fraction of variant appears to decrease and to increase again in January even when cases are decreasing rapidly. This can occur if samples are increasingly weighted towards older, unidentifiable, cases.

**Figure 8.**
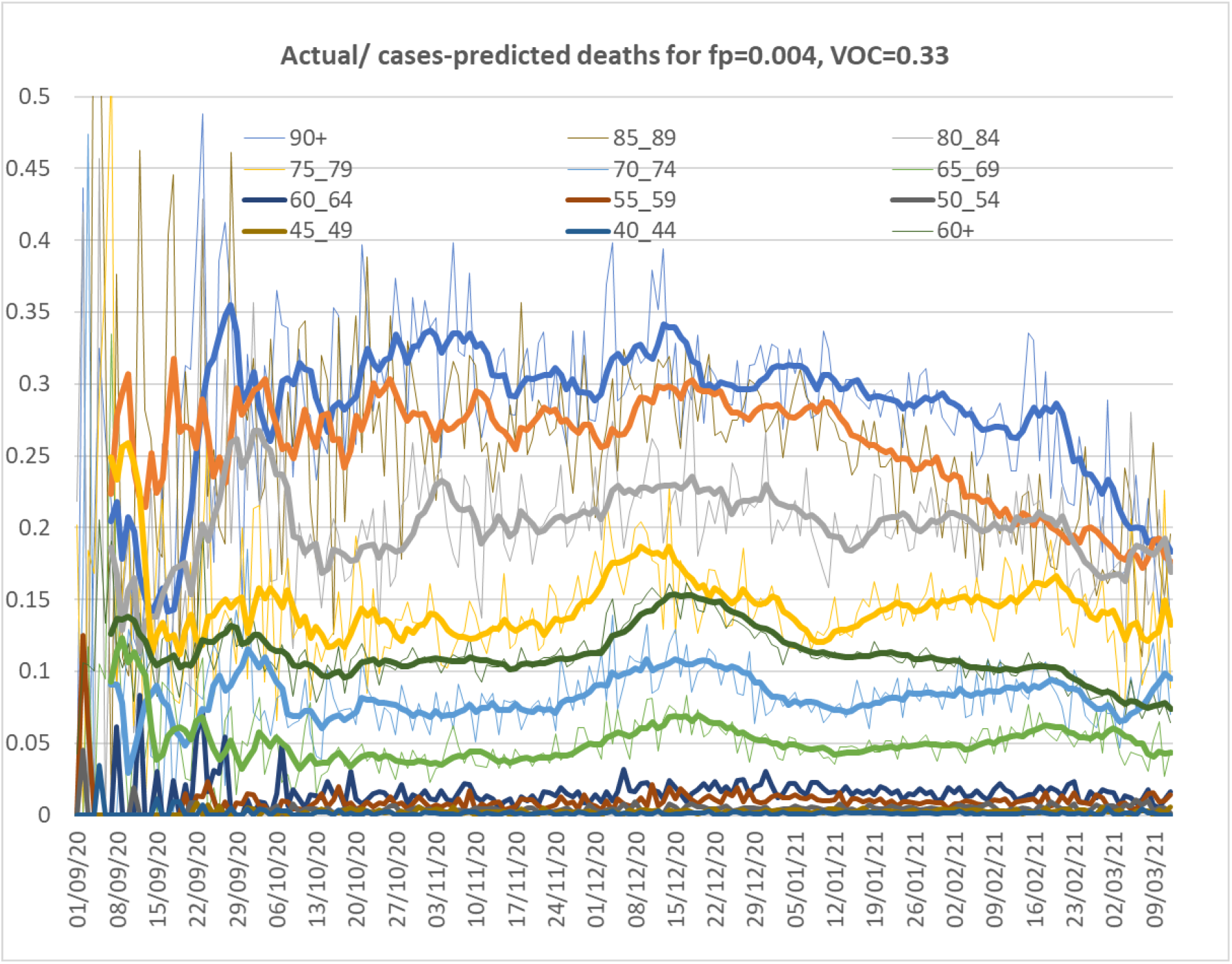
Ratio of actual deaths to deaths predicted by cases, as in Fig 4, but allowing for false positives and VOC, using the 28-day gamma distribution of time from first positive test to death. Case and deaths data from .gov.uk, VoC data from Sanger, the thicker lines are for 7-day rolling averages, to smooth out the daily variance in the fainter lines which give some indication of the statistical uncertainty. A perfect model would give time-independent values for each age group: the only significant deviations come from the unexplained excess in deaths in mid December, and a systematic drop from January onwards.

Inclusion of the effect of VOC, still leaves a discrepancy during December, because the growth of the VOC in ONS data between late November and early December is too late to match deaths in December. Such imperfect correlation suggests that other causes of higher than expected deaths, such as seasonal effects and hospital pressures played a role. Pressures on staff and resources in hospitals resulting from high occupancy have been shown to impact outcomes [25,26] and it would be surprising if this were not the case during the second wave of Covid-19 in England. Figure 11 shows that after peaking during November, both admissions and number of ICU beds occupied declined, and did not reach the same value as they had at their peaks until some two weeks into December, by which time cases were declining, and the surge in CFRs was already under way. We cannot rule out the effect of cumulative stress on hospital resources, particularly staff, but the data do not show a conclusive association.

### 4.3 Evidence for effects of vaccination

Figures 3-5 already suggested deaths may be falling more than the number of cases during February and March. We have already introduced corrections for false positive and VOC, and in Fig 8 we revisit the analysis from Fig, 4, with these corrections in place.

Figure 8 suggests that, even after accounting for B.1.1.7 and false-positives, actual/ cases-predicted deaths began to reduce from mid January, firstly in the oldest age groups, and subsequently in progressively younger groups. It is reasonable to interpret this reduction as a result of vaccination of the earlier vaccination of these groups[27]. Protection from the vaccine is anticipated to take some two weeks or more to be effective, and reduced serious infection will take at least a week to show significantly in a reduction in deaths. By the end of December more than 500,000 people aged 80 and over, some 15% of that age group, had received a first vaccination and by 10 January that had risen to 40%.

It is worth recalling that the CFR should be independent of the amount of infection. So the CFR drop which is evident here is in addition to the reduction in deaths due to immunity. An explanation for the CFR in terms of vaccine requires that the benefits are not only the 70-90% drop in infectivity revealed by the pharmaceutical trials, but an additional benefit of coming from those vaccinees who become infected suffer a milder form of COVID.

Some further words of caution are important: From mid-January to mid-March, cases were falling rapidly, for example by a factor of more than 4 during February, so a large proportion of deaths will come from the tail of the distribution, which in turn is sensitive to the choice of function (Fig 2). So while the vaccine effect is qualitatively evident, it is not realistic for us at this point to fit or to make quantitative statements about the impact of vaccination based on epidemiology. A crucial missing piece of data is the fraction of vaccinated patients among those recorded COVID deaths. This has not been made public, but since vaccine uptake has been reported at 90% it is likely to be significant, and could be easily and reliably determined from clinical data.

## 5 Discussion and Conclusions

We have built on our January report [1] which first revealed the December surge in case fatality ratio, now conclusively accepted as evidence of higher lethality of B.1.1.7. The WSS model from that paper has been improved by implementing gamma distributions for the time “shift” between positive test and death, and by generating UK-based estimates of age-weighted mortality. The case-fatality ratio is a key feature of the model which is largely independent of case numbers, thus changes in the CFR reveal changes to the lethality of the virus itself.

The CFR appears to be low for September 2020. At this stage, cases were low, and the statistics are noisy, but we show that discrepancy can be removed by assuming a modest 0.4% false positive rate.

A marked rise in the CFR is correlated with the spread of the B.1.1.7 variant. This is evident in national level data, as we previously reported [1], and in regional data: specifically, the rise in CFR occurs later in those regions where B.1.1.7 became dominant later. This implies that the B.1.1.7 variant is approximately 33% more deadly, in addition to being around 50% more infectious. This effect is, regrettably, now playing out across Europe where many countries are only now putting measures into place to deal with the threat.

A sharp and ongoing drop in the CFR is evident on the data from January 2021, age group analysis shows it to occur earliest and most strongly in the oldest age groups. We find no dependence on region: in all areas the decrease is similar. The correlation with the vaccination roll-out programme suggests that this is a plausible causal factor. If so, preliminary data for the near-fully vaccinated age groups implies that vaccination confers about 50% lower risk of death, in addition to the well-established 70-90% protection from infection. There are several possible confounding factors in our epidemiological study, but such a conclusion should be easier to obtain from clinical studies with confidential data.

Finally, we note that this work is based only on publicly-available data. Other UK data may exist which would enable precise association of deaths with vaccination and variant status.

However, delays and difficulty obtaining this data would make our analysis obsolete before publication in an epidemic situation. Our WSS model is, by design, simple enough to apply using publicly-available data anywhere in the world. We have shown its main value comes from its ability to detect novel phases of the epidemic immediately they appear, by *failing* to reproduce the data.

A key assumption behind our analysis is that the parameters and sensitivities of PCR tests have been constant since the beginning of September. This requirement also underpins the assumption that the fraction of false positive tests has been constant since that time. The issue is emphasized in the UK Government release during October 2020 [13].

We use data for deaths within 28 days of the date of first positive tests, so the modelled CFRs are specific to this case, excluding parts the gamma distribution implying shifts beyond 28 days. For example, we would expect CFRs for deaths within 60 days to be roughly 10% larger.

## Data Availability

All data is publicly available online via links provided in the paper. Code is available on request.

## Contributors

DJW designed the analysis for the project, and he and JAA did parallel computations in excel and R. GJA focused on issues concerning VOC B.1.1.7 prevalence and its interpretation. All authors contributed to the writing of the paper. We are very grateful to David Spiegelhalter for feedback and comments.

## Patients and Public statement

Patients or the public were not involved in the design, or conduct, or reporting, or dissemination plans of our research.

## Ethical approval

No ethical approval was required for this research.

## Data sharing

All data used in this project is available in public domain as referenced

## Transparency

The lead author affirms that the manuscript is an honest, accurate, and transparent account of the study being reported; that no important aspects of the study have been omitted; and that any discrepancies from the study as planned have been explained.

## Dissemination to participants

Since this research uses public demographic data for the whole of the UK, there are no plans for dissemination of this research to specific participants, beyond publishing it.

